# A clinical decision making model for NTSV deliveries

**DOI:** 10.64898/2026.02.24.26346894

**Authors:** Lauren B. Crabtree, Martin Frasch, Ciprian P Gheorghe

**Affiliations:** Department of Gynecology and Obstetrics, Loma Linda School of Medicine, Loma Linda, California, USA; Department of Obstetrics and Gynecology, Institute on Human Development and Disability, University of Washington School of Medicine, Seattle, Washington, USA

## Abstract

**Objective:** To evaluate modifiable antepartum and intrapartum factors associated with nulliparous, term, singleton, vertex (NTSV) cesarean delivery and to model risk stratified induction timing strategies that minimize cesarean risk across maternal risk profiles.

**Study Design:** This retrospective cohort study included all NTSV deliveries at a tertiary care center from January 2015 through August 2025 (overall cohort n=10,525; limited risk cohort n=5,663).

Machine learning identified key predictors of cesarean delivery, with maternal age and pre pregnancy body mass index (BMI) used to define low, moderate, and high risk strata.

Logistic regression estimated the association between induction and cesarean delivery, and a Monte Carlo simulation compared elective induction at 39, 40, or 41 weeks versus expectant management to 42 weeks within each stratum.

**Results:** Cesarean delivery occurred in 20.1% of the overall cohort and 19.0% of the limited risk cohort, with a U shaped relationship between gestational age and cesarean risk and lowest rates at 38-39 weeks.

Induction was associated with higher cesarean rates than spontaneous labor in both cohorts (overall: 24.1% vs. 17.1%; limited risk: 22.9% vs. 15.7%) after adjustment for age, BMI, and gestational age.

No single induction policy minimized cesarean risk across all strata.

For high risk patients (age >=35 years and BMI >=35), induction at 39 weeks yielded the lowest modeled cesarean rate, whereas later delivery (40 to 41 weeks or expectant management to 41 weeks) was favored for low and moderate risk patients.

A universal 39 week induction policy for low and moderate risk strata modestly increased modeled cesarean rates, adding an estimated 46 cesarean deliveries.

**Conclusion:** Gestational age at delivery and induction strategy are key modifiable determinants of NTSV cesarean delivery, but optimal timing varies by maternal age and BMI risk profile, supporting risk stratified rather than universal 39 week induction policies.

## INTRODUCTION

Cesarean delivery has become a core metric for assessing the quality and safety of obstetric care, with particular emphasis on limiting primary cesarean birth among nulliparous, term, singleton, vertex (NTSV) pregnancies.^1–2^ National initiatives, including the US Department of Health and Human Services’ Healthy People 2030 goal to reduce NTSV cesarean rates to 23.6%, underscore the importance of minimizing unindicated cesarean delivery while ensuring timely access to when needed.^3–4^ Although cesarean delivery can be life-saving, it carries increased risks for maternal morbidity when compared to vaginal delivery (e.g., hemorrhage, infection, thromboembolic events, and prolonged recovery) as well as the risk for complications in subsequent pregnancies. Therefore, NTSV cesarean rate has been widely adopted as a key obstetric quality indicator because it reflects clinical decision-making in a generally low-risk population and is directly influenced by antepartum counseling and intrapartum management strategies.

However, the demographic and clinical profile of pregnant patients in the United States has changed markedly since the introduction of cesarean-related quality benchmarks. Currently, more than half of patients enter pregnancy overweight or obese and a growing proportion are aged 35 years or older.^5–6^ These trends have raised concerns about the applicability of uniform cesarean delivery benchmarks to increasingly heterogeneous obstetric populations. As maternal risk profiles have evolved, some have argued that cesarean reduction initiatives insufficiently incorporate patient-level risk adjustment in labor management strategies and may inadvertently increase maternal and neonatal morbidity by promoting prolonged labor, particularly during the second stage, in attempts to avoid cesarean delivery.^7–11^ Accordingly, there is a pressing need to identify modifiable factors associated with cesarean delivery to inform individualized labor management strategies that balance safety and quality across diverse clinical contexts.

Evidence from recent clinical trials and population-based studies highlights the potential for targeted intrapartum interventions to influence cesarean rates.^12–16^ The ARRIVE Trial demonstrated that, among low-risk nulliparous women, elective induction of labor at 39 weeks was associated with a lower cesarean rate compared with expectant management, without an increase in adverse perinatal outcomes.^17^ Subsequent population-based analyses have suggested that dissemination of ARRIVE was followed by increases in 39-week induction and modest declines in cesarean delivery among low-risk nulliparous patients in the United States, supporting the potential for structured induction strategies as a mechanism to reduce cesarean births.^18^ These data underscore the importance of delineating which antepartum and intrapartum factors are amenable to modification and how such factors influence NTSV cesarean rates in real-world practice.

In this context, the present study aimed to examine antepartum and intrapartum factors associated with NTSV cesarean delivery in a large-scale, risk-stratified data set to optimize patient management and labor/induction polices aligned with the objective of safely hitting NTSV cesarean goals in a more medically complex population. To achieve this goal, we conducted a retrospective cohort study including all eligible NTSV deliveries at Loma Linda Children’s Hospital between January 2015 and August 2025, with a focus on identifying determinants that may be amenable to modification during the antepartum and intrapartum periods.

## MATERIALS AND METHODS

### Study Design and Setting

This retrospective cohort study analyzed all NTSV deliveries from January 2015 through August 2025 at Loma Linda Children’s Hospital. The study protocol was reviewed and approved by the Loma Linda Children’s Hospital Institutional Review Board, which granted a waiver of informed consent for secondary analysis of existing and de-identified data.

### Study Population

Two cohorts were established to evaluate the generalizability and robustness of study findings. First, the overall cohort included all NTSV deliveries at 37- to 42-weeks gestation (N=10,525). A second sensitivity cohort, henceforth referred to as the limited-risk cohort, was defined as 39-weeks gestation and above and excluded pregnancies/deliveries complicated by diabetes, pre-existing chronic hypertension, and premature rupture of membranes (PROM; n=5,663). This second cohort was used to assess the robustness of delivery timing effects following the exclusion of common high-risk conditions and medical indications for induction and delivery before 39-weeks gestation. The NTSV population was defined according to Joint Commission specifications.^2^

### Data Source and Variables

#### Data Source

We analyzed deliveries at Loma Linda Children’s Hospital between January 2015 and August 2025 using data from the institutional perinatal database, an actively maintained repository derived from electronic health records and delivery logs. The database prospectively captures comprehensive maternal and neonatal information for all deliveries, with all personally identifiable data removed. The analytic cohort was limited to NTSV deliveries, and we further reduced the dataset from 10,661 to 10,525 after removing patients with missing records.

#### Variable Selection

Seven base features were selected for analytic modeling based on completeness, clinical relevance, and significance in prior obstetric literature: maternal age, pre-pregnancy body mass index (BMI), gestational age at delivery, birth weight, labor induction, race/ethnicity, and insurance payer type.

#### Data Processing and Machine Learning

We restricted our analysis to records with complete data. To identify key modifiable and non-modifiable predictors of cesarean delivery, we conducted machine learning analysis. Candidate predictors included maternal age, pre-pregnancy body mass index (BMI), gestational age at delivery, labor induction status, race and ethnicity, insurance status, and parity. To assess the robustness of predictor selection, we conducted identical analyses on both the overall cohort (n=10,525) and a sensitivity (limited-risk) cohort (n=5,663), allowing assessment of predictor consistency across cohorts with different risk profiles.

### Risk-Stratified Analysis of Delivery Management

Our machine learning analysis established that among the key predictors of cesarean delivery in NTSV pregnancies, only gestational age at delivery and labor induction decisions are modifiable during the antenatal and intrapartum periods. The most influential nonmodifiable predictors identified by the model, maternal age and pre-pregnancy BMI, were used to define risk strata within both the overall and limited-risk cohorts. Risk-stratified analyses were then conducted to examine the associations between these inherent maternal characteristics and the optimal timing of delivery among women with spontaneous and induced labor.

#### Statistical Analysis

Categorical variables were compared across binary outcomes using Pearson’s chi-square or Fisher’s Exact tests (*p*<0.05), and continuous variables were compared using Welch’s t tests (*p*< 0.05). Logistic regression, adjusted for gestational age, maternal age, and BMI, evaluated the association between induction and cesarean delivery, reported as odds ratios (OR) with 95% confidence intervals. To characterize nonlinear and interaction effects among predictors, a Random Forest classifier (100 trees, maximum depth of 5) was used to estimate variable importance. Analyses were performed in Python (version 3.10.16) using pandas, NumPy, SciPy, statsmodels, scikit-learn, Matplotlib, and Seaborn.

### Monte Carlo Simulation

A Monte Carlo simulation was developed to build a clinical decision-making model for timing of induction of labor across maternal risk groups. The model evaluated four management strategies within each of the previously-described risk groups: elective induction at 39, 40, or 41 weeks, and expectant management until 42 weeks. For each strategy and risk group, we generated 10,000 “virtual patients” whose trajectories were simulated with the primary outcome being the probability of cesarean delivery under each induction-timing strategy to inform a risk-stratified clinical decision model. Competing risks were explicitly modeled, including spontaneous labor at any gestational age, development of new maternal complications prompting delivery, and cesarean delivery at the time of induction or spontaneous labor.

## RESULTS

### Cohort Characteristics

A total of 10,661 deliveries were identified, of which 10,525 met the study criteria and were included in the overall cohort (N= 10,525). After applying sensitivity analysis exclusions, 5,663 deliveries comprised the limited-risk cohort (over 39 weeks and no diabetes, pre-existing chronic hypertension, or PROM; n= 5,663). The overall cohort, in contrast with the limited-risk cohort, included early-term deliveries at 37 and 38 weeks and patients with diabetes, pre-existing hypertensive disorders, and/or PROM. Compared with the overall cohort, patients in the limited-risk cohort were younger (26.7 vs 27.1 years, *p* < 0.001) and had lower mean pre-pregnancy BMI (27.2 vs 27.7, *p* < 0.001). Baseline maternal characteristics and delivery outcomes are presented in Table 1.

**Table 1.** Baseline characteristics and outcomes of the Overall and Sensitivity cohorts.

| **Characteristic** | **Overall cohort**<br>**(N=10,525)** | **Sensitivity cohort**<br>**(N=5,664)□** | ***P* value** |
| --- | --- | --- | --- |
| Maternal Age (SD) | 27.1 (5.7) | 26.7 (5.6) | <0.001 |
| Pre-pregnancy BMI (SD) | 27.7 (7.0) | 27.2 (6.4) | <0.001 |
| Gestational Age at Delivery (SD) | 39.2 (1.1) | 39.9 (0.7) | <0.001 |
| Induction of Labor (%) | 5,848 (55.6) | 3,149 (55.6) | 0.956 |
| Cesarean Delivery (%) | 2,118 (20.1) | 1,078 (19.0) | 0.003 |

□ Sensitivity cohort excludes patients with preexisting hypertension, diabetes, or premature rupture of membranes.

The cesarean delivery rate was lower in the limited-risk cohort compared with the overall cohort (19.0% vs 20.1%; *p*= 0.003), despite equivalent frequency of labor induction (55.6% [3,149] vs 55.6% [5,848]; *p=* 0.956). Gestational age distribution varied significantly between cohorts, as by definition the low-risk cohort excluded deliveries before 39 weeks and had a higher mean gestational age compared with the overall cohort (39.9 ± 0.7 vs 39.2 ± 1.1 weeks, p<0.001).

### Gestational Age and Cesarean Delivery Risk

#### Overall Cohort Analysis

Cesarean delivery rates varied by gestational age in a U-shaped pattern, with the lowest rates occurring at 38-39 weeks gestation, and higher rates observed at both earlier and later gestational ages. Cesarean delivery occurred in 24.8% of deliveries at 37 weeks, decreasing substantially to 19.2% and 19.7% at 38 and 39 weeks, respectively. At later gestational ages, the cesarean rate was 20.5%, 23.8%, and 62.0% at 40, 41, and 42 weeks, respectively.

#### Limited-Risk Cohort Analysis

In the limited-risk cohort, which excluded pregnancies with indications for delivery prior to 39 weeks, cesarean delivery rates were lowest at 39 weeks (18.45%; Table 2). Beyond 39 weeks, cesarean rates increased in a manner similar to the primary cohort, increasing to 19.86%, 22.81%, and 62.86% at 40, 41, and 42 weeks, respectively.

**Table 2.** Cesarean Section Rates by Gestational Age and Labor Onset.

| **Gestational Age** | **Overall CS% (n)** | **Limited-Risk CS% (n)** | **P value** |
| --- | --- | --- | --- |
| *Spontaneous Labor Onset* |  |  |  |
| 39 weeks | 14.8% (257) | 14.8% (199) | 1.000 |
| 40 weeks | 15.3% (178) | 14.2% (140) | 0.542 |
| 41 weeks | 20.7% (42) | 19.4% (33) | 0.796 |
| 42 weeks | 52.0% (13) | 42.9% (6) | 0.741 |
| *Induced Labor Onset* |  |  |  |
| 39 weeks | 22.2% (474) | 20.2% (322) | 0.146 |
| 40 weeks | 23.8% (304) | 23.5% (258) | 0.885 |
| 41 weeks | 24.9% (114) | 23.9% (104) | 0.755 |
| 42 weeks | 69.2% (18) | 72.7% (16) | 1.000 |

### Machine Learning Identified Risk Factors

The machine learning model identified maternal age, pre-pregnancy BMI, gestational age, and labor induction as key predictors of cesarean section. Among these, maternal age and pre-pregnancy BMI were the most influential nonmodifiable predictors and were used to define patient strata for risk-stratified analyses to examine associations between these inherent maternal characteristics and the optimal timing and mode of delivery, focusing on gestational age and induction of labor as actionable clinical factors.

### Labor Induction and Cesarean Delivery Risk

#### Overall Cohort Analysis

As shown in Table 2, cesarean delivery rates were higher among induced compared to spontaneous labor across all gestational ages (24.1% vs 17.1%; *p*<.001). In adjusted analyses controlling for gestational age, maternal age, and BMI, labor induction increased the odds of cesarean delivery by 28% (OR= 1.06; *p*<.001).

#### Limited-Risk Cohort Analysis

In the limited-risk cohort (Table 2), induction of labor remained associated with a higher cesarean delivery rate compared with spontaneous labor (22.93% vs 15.67%; *p*<.001). After adjustment, induction was associated with increased odds of cesarean delivery (adjusted OR, 1.38; *p*<.001).

### Risk-Stratified Analysis of Delivery Management

To account for risks associated with inalterable maternal characteristics, we created a risk stratification schema based on maternal age and BMI, which our machine learning model identified as the most significant inalterable risk factors for cesarean delivery, to identify the optimal delivery timing strategy with the least risk for cesarean delivery. The low-risk group was defined as younger than 35 years with BMI <30. The moderate-risk group included patients who met one, but not both, elevated risk criterion: either advanced maternal age (≥35 years) or BMI 30.0–34.9. The high-risk group was defined by the coexistence of advanced maternal age (≥35 years) and BMI ≥35.

#### Overall Cohort Results

In the overall cohort, risk stratification classified 6,569 (62.4%) patients as low risk, 3,565 (33.9%) as moderate risk, and 391 (3.7%) as high risk. The lowest cesarean delivery rates were not consistent across risk strata for any singular gestational age for induced or spontaneous labor. Among low risk patients, the lowest cesarean rate for spontaneous labor occurred at 38 weeks (9.5%) and for induced labor at 37 weeks (13.8%). In the moderate risk group, the lowest cesarean rate for spontaneous labor occurred at 39 weeks (20.4%) and for induced labor at 37 weeks (31.5%). In the high risk group, cesarean rates for spontaneous labor were highest at 37 weeks (80.0%) and lower at later gestational ages (35.5% at 38 weeks, 52.9% at 39 weeks, 40.0% at 40 weeks, and 0.0% at 41 weeks), whereas cesarean rates for induced labor were lowest at 41 weeks (29.4%) compared with 41.9% at 37 weeks, 46.0% at 38 weeks, 31.5% at 39 weeks, and 37.2% at 40 weeks. See Table 3.

**Table 3.** Overall Cohort Cesarean Delivery Rates by Gestational Age and Risk Level.

| **Gestational Age** | **Low Risk** | **Medium Risk** | **High Risk** |
| --- | --- | --- | --- |
| *Spontaneous Labor Onset* |  |  |  |
| 37 Weeks | 20.1% (328) | 40.9% (171) | 80.0% (15) |
| 38 Weeks | 9.5% (714) | 21.1% (285) | 35.5% (31) |
| 39 Weeks | 12.5% (1,209) | 20.4% (495) | 52.9% (34) |
| 40 Weeks | 12.5% (843) | 24.2% (314) | 40.0% (10) |
| 41 Weeks | 15.3% (131) | 34.8% (66) | 0.0%* (6) |
| *Induced Labor Onset* |  |  |  |
| 37 Weeks | 13.8% (537) | 31.5% (406) | 41.9% (62) |
| 38 Weeks | 16.9% (498) | 33.3% (384) | 46.0% (63) |
| 39 Weeks | 16.9% (1,214) | 31.1% (817) | 31.5% (108) |
| 40 Weeks | 21.0% (785) | 29.9% (448) | 37.2% (43) |
| 41 Weeks | 20.4% (284) | 32.7% (156) | 29.4% (17) |

Data presented as rate % (patient count).

#### Limited-Risk Cohort Results

The limited-risk cohort was likewise stratified by risk based on our criteria, with 3,731 (65.88%) patients classified as low-risk, 1,783 (31.49%) classified as moderate-risk, and 149 (2.63%) classified as high-risk. Among patients in the low- and moderate-risk strata, induction of labor was associated with higher cesarean delivery rates across all gestational ages. Among low-risk patients, the lowest cesarean delivery rate occurred with spontaneous labor at 40 weeks (12.3%), whereas the lowest rate following induction was observed at 39 weeks (13.1%). For moderate-risk patients, the lowest cesarean rates were observed at 39 weeks for both spontaneous labor (19.7%) and induction (22.5%). In contrast, among high-risk patients at 39 weeks, cesarean delivery was more common among spontaneous labor (48.0%) than induction of labor (31.9%); however, by 40 weeks, this trend reversed. See Table 4.

**Table 4.** Limited-Risk Cohort Cesarean Delivery Rates by Gestational Age and Risk Level.

| **Gestational Age** | **Low Risk** | **Medium Risk** | **High Risk** |
| --- | --- | --- | --- |
| *Spontaneous Labor Onset* |  |  |  |
| 39 Weeks | 13.1% (1,054) | 19.7% (447) | 48.0% (25) |
| 40 Weeks | 12.3% (733) | 22.5% (285) | 28.6% (7) |
| 41 Weeks | 15.9% (113) | 29.6% (54) | 0.0%* (5) |
| 42 Weeks | 60.0% (5) | 20.0% (5) | No Data |
| *Induced Labor Onset* |  |  |  |
| 39 Weeks | 16.6% (1,087) | 28.3% (743) | 31.9% (91) |
| 40 Weeks | 20.7% (686) | 29.7% (407) | 36.4% (33) |
| 41 Weeks | 19.3% (249) | 31.8% (142) | 29.4% (17) |
| 42 Weeks | 77.8% (9) | 75.0% (4) | No Data |

Data presented as rate % (patient count). \*Small sample size may affect rate reliability.

### Monte Carlo Simulation

Given that no single universal induction strategy (e.g., elective induction of all patients at 39 weeks) optimized cesarean outcomes in our cohort, we constructed a Monte Carlo simulation model to estimate induction strategies tailored to patient characteristics that minimized modeled cesarean risk. Across all scenarios, the simulation confirmed that no single, universal induction policy minimized cesarean delivery for all patients. Instead, the optimal timing of delivery varied by baseline risk at admission. In the high-risk group, induction at 39 weeks produced the lowest cesarean rate at 30.8%, which rose significantly to 39.5% when induction was postponed to 40 weeks. In contrast, cesarean delivery rates in the moderate-risk group differed significantly across timing strategies, with higher cesarean rates for induction at 39 weeks (28.15%; *p*<0.01) compared with 40 weeks (27.47%; *p*<0.01) and 41 weeks (27.36%; *p*<0.01). Although the absolute differences are modest among the moderate-risk patient group, they may still be clinically meaningful when projected across large populations. Expectant management until 41 weeks was similarly favored for patients in the low-risk strata, with later gestational age at delivery being associated with a significantly lower risk for cesarean delivery: 16.45% (*p*<0.001) at 39 weeks, 18.90% (*p*<0.001) at 40 weeks, and 15.75% (*p*<0.001) at 41 weeks.

#### Policy Simulation: Mandatory 39-week Induction

We then examined the impact of a hypothetical policy mandating elective induction at 39 weeks among low- and medium-risk patients, as the ARRIVE Trial suggested might be optimal.^17^ Relative to the simulation-identified optimal strategy of expectant management until 41 weeks, a blanket 39-week induction policy increased the projected number of cesarean deliveries. In the low-risk cohort, the cesarean rate rose from 15.7% under the optimal strategy to 16.5% (*p*<0.00001) with universal 39-week induction, corresponding to approximately 30 additional cesarean deliveries per 3,731 patients. In the medium-risk cohort, the cesarean rate increased from 27.3% to 28.2% *(p<*0.00001), yielding an estimated 16 additional cesarean deliveries per 1,783 patients. Taken together, universal 39-week induction in these two groups was associated with roughly 46 excess cesarean deliveries in the modeled cohort, while converting a substantial proportion of spontaneous labors to scheduled inductions. These findings suggest that among low- and moderate-risk patients earlier induction increases the risk for cesarean delivery, whereas deferring delivery to 41 weeks preserves or modestly improves cesarean outcomes. However, high-risk patients do benefit from earlier delivery timing.

## DISCUSSION

In this analysis of 10,525 NTSV deliveries, optimizing delivery timing emerged as an actionable strategy during the antenatal and intrapartum periods. Our analysis identified gestational age at delivery and labor induction strategies as the principle modifiable variables during the antenatal and intrapartum periods. Cesarean rates demonstrated a U shaped relationship with gestational age, with relatively lower rates around the 38-39 weeks and higher rates at both earlier and later gestations. However, the gestational age associated with the lowest risk was not consistent across all risk strata and varied by baseline risk level and mode of labor onset. Maternal age and BMI emerged as the strongest non modifiable predictors of cesarean delivery, and induction of labor was consistently associated with higher cesarean rates than spontaneous labor, even after adjustment for maternal age, BMI, and gestational age.

Our Monte Carlo simulation further underscored that the impact of elective induction on cesarean risk is highly context dependent, with different timing strategies favored across low□, medium□, and high risk groups rather than a single, universally optimal gestational age as proposed in the ARRIVE Trial.^17^ Among patients at highest baseline risk, earlier planned delivery was favored in the model, whereas in medium□ and low□risk patients later induction or continued expectant management yielded lower cesarean rates. A hypothetical universal 39-week induction policy for low□ and medium□risk patients increased the projected number of cesarean deliveries and converted a substantial proportion of spontaneous labors into scheduled inductions, highlighting the limitations of unstratified, one□size□fits□all approaches. Taken together, these findings support incorporation of maternal factors, most notably age and pre-pregnancy BMI, into counseling and institutional protocols regarding timing of delivery, and argue against the adoption of a universal induction protocol as a primary cesarean reduction strategy.

### Strengths and Limitations

Strengths include rigorous data quality assessment, individualized risk stratification by maternal age and BMI, and real-world applicability within an unselected tertiary population. Limitations include a single-center retrospective design and small sample sizes at gestational extremes. The absence of more granular clinical data and neonatal outcomes limits generalization.

### Future Directions

Future studies should validate these findings in prospective, multi-center studies, which would allow for better generalization and more precise risk stratification. Further, the development and validation of individualized risk prediction models may facilitate not only improved personalized counseling and shared decision-making but also inform institutional guidelines and policy efforts aimed at optimizing timing of delivery.

## Data Availability

All data produced in the present study are available upon reasonable request to the authors

## ACKNOWLEDGEMENTS

We thank the perinatal data registry staff at Loma Linda University Medical Center for their assistance with data collection and quality assurance.

